# Safety, Pharmacokinetics, Biomarker Response, and Efficacy of E6742, a Dual Antagonist of Toll-Like Receptors 7 and 8, in a First-in-Patient, Randomized, Double-Blind, Phase 1/2 Study in Systemic Lupus Erythematosus

**DOI:** 10.1101/2024.04.26.24306410

**Authors:** Yoshiya Tanaka, Atsushi Kumanogoh, Tatsuya Atsumi, Tomonori Ishii, Fumitoshi Tago, Mari Aoki, Shintaro Yamamuro, Shizuo Akira

## Abstract

**Objectives:** To evaluate the safety, tolerability, pharmacokinetics (PK), biomarker response, and efficacy of E6742 in a phase 1/2 study in patients with systemic lupus erythematosus (SLE).

**Methods:** Two sequential cohorts of SLE patients were enrolled and randomized to 12 weeks of twice-daily treatment with E6742 (100 or 200 mg; n = 8 or 9) or placebo (n = 9).

**Results:** The proportion of patients with any treatment-emergent adverse events (TEAEs) was 58.8% in the E6742 group (37.5% for 100 mg; 77.8% for 200 mg) and 66.7% in the placebo group. No Common Terminology Criteria for Adverse Events ≥ Grade 3 TEAEs occurred. PK parameter levels were similar between SLE patients and healthy adults in previous phase 1 studies. The interferon gene signature (IGS) and levels of proinflammatory cytokines (interleukin-1β, interleukin-6, tumor necrosis factor-α) after ex-vivo challenge with a Toll-like receptor 7/8 agonist were immediately decreased by E6742 treatment. Dose-dependent improvements in the British Isles Lupus Assessment Group-based Composite Lupus Assessment response were observed at Week 12 in the E6742 (37.5% for 100 mg; 57.1% for 200 mg) and placebo (33.3%) groups. E6742 also had therapeutic effects on other symptoms, including skin inflammation, arthritis, and levels of anti-double-stranded DNA antibodies and complements.

**Conclusions:** E6742 had a favorable safety profile and was well tolerated, with marked IGS responses and sufficient efficacy signals in patients with SLE. These results provide the first clinical evidence to support E6742 in the treatment of SLE, and support larger, longer-term clinical trials.

**Trial registration number:** NCT05278663.

**KEY MESSAGES:** **What is already known on this topic**

- Because of the limited efficacy and safety concerns of current drug therapies, unmet medical needs remain for many patients with systemic lupus erythematosus (SLE), necessitating new, more efficacious drugs.
- There is strong evidence for the relationship between Toll-like receptor (TLR)7/8 and SLE pathophysiology, and two phase 1 clinical studies of E6742, a small molecular selective dual antagonist of TLR7/8, in healthy adults showed good tolerance without safety issues.

**What this study adds**

- E6742 was well tolerated in this phase 1/2 clinical trial of patients with SLE, demonstrating a favorable safety profile and providing a markedly improved interferon gene signature and sufficient efficacy signals.

**How this study might affect research, practice or policy**

- This study provides the first clinical evidence to suggest that E6742, as a first-in-class TLR7/8 inhibitor, may be beneficial for SLE.
- The study outcomes also support larger, longer-term clinical trials of E6742.

## INTRODUCTION

Systemic lupus erythematosus (SLE) is a heterogeneous autoimmune disease characterized by systemic inflammation in various organs, including the skin, joints, kidneys, and central nervous system. SLE is more common in young female adults (9:1 ratio of females to males) [1]. The reported global prevalence of SLE varies considerably, ranging from 3.2 to 517.5 per 100,000 individuals [2, 3]. According to the European Alliance of Associations for Rheumatology (EULAR) recommendations for the management of SLE (2023 update), the treatment goal is to achieve remission or low disease activity [4]. Hydroxychloroquine (HCQ) is currently recommended for all SLE patients unless contraindicated, while the use of glucocorticoids has shifted from mainstay of treatment to a “bridging therapy” limited to minimal use on the basis of detrimental effects. In patients who do not respond to HCQ (alone or in combination with glucocorticoids) or are unable to reduce glucocorticoids, addition of immunosuppressants and/or biologics should be considered. However, most of these conventional drugs cause serious side effects, leading to the anticipation of development of more targeted therapies [5–7].

While the adaptive immune response, specifically involving B cells and autoantibodies, has traditionally been considered a central component of SLE pathogenesis, recent evidence and genome-wide association studies have highlighted the importance of the innate immune response in disease development and progression [5]. Biologics have been approved for SLE target molecules produced by the innate immune system, namely type I interferon (IFN) and the B lymphocyte stimulator pathway, which play key roles in the activation of the adaptive immune response in SLE. Therefore, the innate immune system has been the focus of new therapeutic targets for SLE. Although the pathophysiology of SLE is not fully understood, there is much evidence indicating that autoimmune responses exerted via type I IFNs and activation of the interferon gene signature (IGS) are important in the pathogenesis. Plasmacytoid dendritic cells (pDCs) are specialized subsets of dendritic cells that produce large amounts of type I IFNs in response to viral or bacterial pathogens. Although these IFNs represent fundamental defense reactions against infections [8–10], their dysregulation can drive autoimmunity. Toll-like receptor (TLR)7, which recognizes single-stranded RNA, is highly expressed on pDCs and is thought to be a major trigger of type I IFN production in SLE [11–14]. There is also strong evidence for a genetic contribution of TLR7 in the pathogenesis of SLE [15, 16]. Compared with TLR7, TLR8 is more abundant in neutrophils, monocytes, and myeloid dendritic cells, inducing strong production of inflammatory cytokines via activation of nuclear factor-κB signaling [17]. Growing evidence has demonstrated that dual inhibition of TLR7/8 may have stronger effects than inhibition of either receptor alone in the treatment of SLE, modulating innate immune responses and suppressing the production of many inflammatory cytokines, including IFN-α. Furthermore, preclinical studies revealed that a dual TLR7/8 inhibitor has the potential to increase the effectiveness of glucocorticoids, indicating a glucocorticoid-sparing potential for TLR7/8 inhibition [18]. E6742 is a selective dual antagonist for TLR7/8 with potential value in resolving the unmet medical needs of SLE patients.

E6742 was evaluated in phase 1 studies testing a single ascending dose (SAD) and a multiple ascending dose (MAD) in healthy adults. The safety results from the SAD and MAD studies demonstrated that E6742 has an adequate safety and tolerability profile [19].

Herein, we present the results of a phase 1/2, double-blind, placebo-controlled study of E6742 to confirm its safety, pharmacokinetics (PK), biomarker response, and efficacy in patients with SLE.

## METHODS

### Study design

This was a randomized, double-blind, placebo-controlled, multicenter, MAD study of E6742 in two sequential cohorts of patients with SLE from 12 study sites in Japan (supplemental list of investigators and study sites). The primary endpoint was to evaluate the safety and tolerability of E6742, the secondary endpoints were to evaluate the PK and IGS after the treatment of E6742. In each cohort, patients were randomized to either E6742 or placebo at a ratio of 2:1 and received multiple oral doses of E6742 or placebo for 12 weeks (85 days). Patients received 100 mg E6742 or placebo twice daily (BID) in Cohort 1 and 200 mg E6742 or placebo BID in Cohort 2. A randomization list was generated by an interactive web response system using a permuted block design. Subjects and all personnel involved with the conduct and the interpretation of the study, including investigators, site personnel, and sponsor staff were blinded to the treatment codes. PK parameters for dose escalation assessment were evaluated by the pre-designated unblinded PK analyst, while all other study personnel remained blinded.

This study consisted of a pre-randomization phase (Day –30 to start of study drug dosing on Day 1), a treatment period (Days 1 to 85), and a follow-up period of 4 weeks (28 days) after the last dose of study drug. The study was registered at ClinicalTrials.gov (<u>NCT05278663</u>). The protocol was approved by the Institutional Review Board for each study site. All patients gave written informed consent before participation. This study was conducted in accordance with the standard operating procedures of the sponsor, which were designed to ensure adherence to the Declaration of Helsinki and Good Clinical Practice.

### Patients

Patients between 18 and 75 years of age who met the 1997 American College of Rheumatology (ACR) criteria, the 2012 Systemic Lupus International Collaborating Clinics (SLICC) criteria, or the 2019 EULAR/ACR criteria for SLE at least 6 months before the informed consent deadline were eligible for enrollment if they met all other entry criteria. These criteria included a requirement for active SLE disease at screening, i.e., exhibited at least one active clinical symptom of SLE (arthritis, rash, myositis, mucosal ulcer, pleurisy, pericarditis, or vasculitis) based on SLE Disease Activity Index 2000 (SLEDAI-2K) assessment. Positive tests for antinuclear autoantibodies, anti-double-stranded DNA (anti-dsDNA) antibodies, and/or anti-Smith (anti-Sm) antibodies were also required, and patients had to be on stable standard SLE therapy for at least 4 weeks prior to the first dose of the study medication. Allowable baseline standard SLE therapies included oral glucocorticoids (≤ 20 mg/day prednisone or equivalent), antimalarials (≤ 400 mg/day HCQ), and immunosuppressants (at least one of the following: ≤ 200 mg/day azathioprine, ≤ 3 g/day mycophenolate, ≤ 15 mg/week methotrexate, ≤ 3 mg/day tacrolimus, and ≤ 200 mg/day ciclosporin).

Patients were excluded if they had any unstable or progressive manifestation of SLE (e.g., active nephritis or active central nervous system involvement) or other inflammatory disease that might confound efficacy assessments. Furthermore, patients were not allowed to have had previous treatment with the following within prespecified timeframes prior to the first dose of the study medication: B-cell targeted therapy, anti-type I IFN antibody, oral Janus kinase inhibitors, B-cell depleting therapy, or immunomodulatory biological therapy.

### Assessments

#### Safety

Safety was evaluated based on all adverse events (AEs) and serious AEs, clinical laboratory parameters (hematology, blood chemistry, urinalysis), vital signs, 12-lead electrocardiogram (ECG), physical examination results, and chest X-rays. AEs were coded using the Medical Dictionary for Regulatory Activities (MedDRA), version 26.0. Severity of an AE was graded on a 5-point scale according to Common Terminology Criteria for Adverse Events (CTCAE v5.0).

#### PK

On Days 1 and 15, plasma samples were collected immediately before administration of the study drug and at 1, 2, 3, and 6 hours post-administration. The plasma concentration of E6742 was measured in patients receiving active treatment using a validated liquid chromatography-tandem mass spectrometry (LC-MS/MS) method.

#### Biomarker response

Blood samples for IGS assessment were collected from all patients at baseline, Weeks 2, 4, 8, and 12, and the follow-up visit. RNA was extracted from whole blood for analysis of IGS expression levels by RNA sequencing using validated methods. Based on previous reports of higher expression of IFN-related genes in SLE patients compared to that in healthy adults [20–22], a total of 127 IGSs were selected for this study (supplemental table S1). Pre- and post-treatment changes in expression of these 127 genes were evaluated, and the most prevalently expressed 21 genes (*EPSTI1*, *HERC5*, *IFI27*, *IFI44*, *IFI44L*, *IFI6*, *IFIT1*, *IFIT3*, *ISG15*, *LAMP3*, *LY6E*, *MX1*, *OAS1*, *OAS2*, *OAS3*, *PLSCR1*, *RSAD2*, *RTP4*, *SIGLEC1*, *SPATS2L*, and *USP18*) were selected for calculation of the “IGS score” [20, 23].

Blood samples for determination of interleukin (IL)-1β, IL-6, and tumor necrosis factor-α (TNF-α) levels after ex-vivo challenge with a TLR7/8 agonist were collected from all patients into heparinized tubes at five time points on Days 1 and 15 (pre-dose, and 1, 2, 3, and 6 hours post-dose). Blood samples were incubated with the dual TLR7/8 agonist R848 (2.4 µM) for 24 hours at 37 [and 5% CO_2_. After centrifugation of the samples, supernatants were collected and stored at −80 □ until the cytokines were measured. Concentrations of cytokines were quantified using the V-PLEX Human Proinflammatory Panel 1 Kit from Meso Scale Discovery, in accordance with the manufacturer’s instructions.

#### Efficacy

The following efficacy outcomes were assessed at Week 12: British Isles Lupus Assessment Group-based Composite Lupus Assessment (BICLA) response, SLEDAI-2K, British Isles Lupus Assessment Group Index 2004 (BILAG-2004), Physician Global Assessment (PGA), Cutaneous Lupus Erythematosus Disease Area and Severity Index (CLASI) activity score, CLASI-50 response (decrease of ≥ 50% from baseline CLASI activity score), tender joint counts (out of 68 joints) and swollen joint counts (out of 66 joints), and serological markers (anti-dsDNA antibody and complement C3/C4 levels). The BICLA response was defined as meeting all of the following criteria: no worsening of total SLEDAI-2K score compared with the baseline level; all BILAG-2004 A scores at baseline improved to B, C, or D, and all BILAG-2004 B scores improved to C or D; no worsening in disease activity to BILAG-2004 A score and no scores ≥ 2 worsening to BILAG-2004 B score compared to baseline level; no worsening in the PGA (an increase of < 0.3 points from baseline); and no discontinuation of trial medication or use of restricted medications beyond the protocol-allowed threshold.

### Statistical analyses

Sample size selection was not based on statistical power. Twelve patients per cohort (8 patients randomized to E6742 and 4 patients randomized to placebo) were considered adequate to evaluate the safety, tolerability, PK, and IGS in patients with SLE. Safety analyses were conducted using the safety analysis set, which included all patients who received at least one dose of the study treatment and had safety data. PK analyses were conducted using the PK analysis set, which included all patients who received at least one dose of the study drug and had sufficient PK data to derive at least one PK parameter. Biomarker analyses were conducted using the full-analysis set, which included all patients who were randomized and received at least one dose of the study treatment and had at least one post-dose efficacy measurement. The 21 most prevalently expressed, IFN-inducible genes were selected as candidate biomarkers for anti-IFN-α therapy in SLE [20]. To assess the level of expression of this gene set, the IGS score for each patient was calculated as the median of the fold change in these 21 genes relative to a pooled healthy adult sample [23]. All efficacy analyses were conducted using the full-analysis set. BICLA response analyses were performed on patients with at least one BILAG-2004 A or BILAG-2004 B score at baseline. Participants who discontinued treatment were imputed as non-responders for all of the visits following treatment discontinuation. Data were analyzed using Statistical Analysis Software (SAS) V.9.4 (SAS Institute, NC, USA).

### Patient and Public Involvement

Patients from the Japanese lupus group were involved to discuss the appropriateness of the trial procedures during the trial. There was no further patient or public involvement in the planning, practice, analysis, or reporting process of the trial.

## RESULTS

### Patient disposition and baseline characteristics

This study was conducted between April 2022 and September 2023. A total of 39 patients were screened, of whom 27 were randomized into the study. Of the 27 randomized patients, 26 received at least one dose of study drug; 9 patients in Cohorts 1 and 2 received placebo, and 17 patients received E6742 at a dosage of 100 mg BID (8 patients in Cohort 1) or 200 mg BID (9 patients in Cohort 2). Most of the treated patients completed the study, with one participant each in the placebo and 200 mg BID groups withdrawing from the study because of lack of efficacy and an AE, respectively (figure 1).

**Figure 1.**
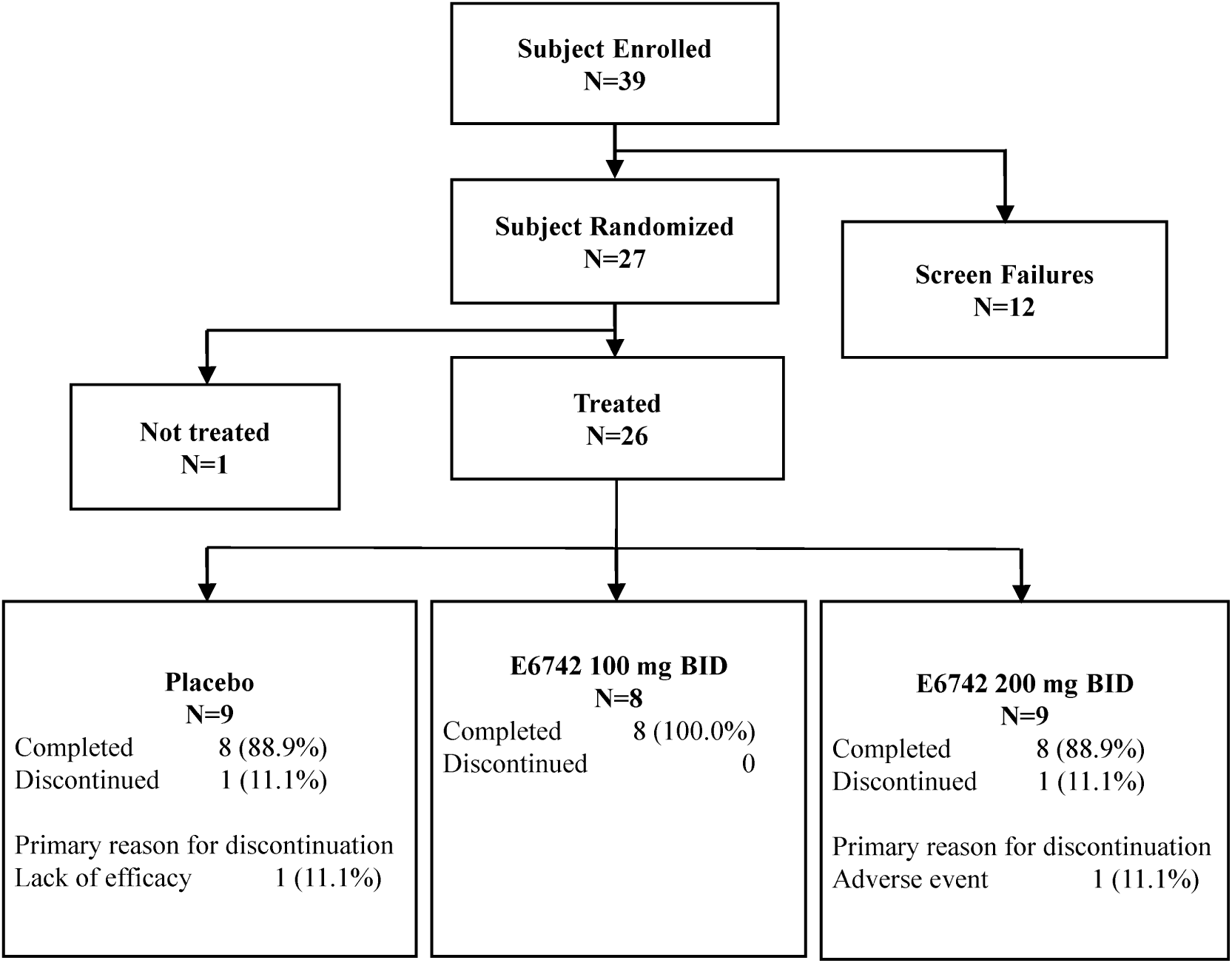
Trial patient disposition. BID, twice daily.

Baseline demographics and disease characteristics are shown in table 1. Although the 100 mg group had a slightly lower mean age and a shorter disease duration than the other groups, the baseline demographics were generally similar across the treatment groups. Regarding clinical characteristics, some disease index parameters, including SLEDAI-2K scores and tender or swollen joint counts, were slightly lower in the 200 mg group than in the other groups. Among the BILAG-2004 components, the three most common manifestations of active SLE were mucocutaneous (80.8%), musculoskeletal (69.2%), and haematological (57.7%). The most frequently used concomitant medication for SLE at baseline was oral glucocorticoid (96.2%), followed by HCQ (80.8%) for all patients. At baseline, 92.3% of the patients were receiving oral glucocorticoid at doses ≤10 mg/day. Overall, at baseline, 34.6% of the patients were taking three medications concomitantly and 53.8% of patients were taking two medications concomitantly for SLE: HCQ, oral glucocorticoid, or an immunosuppressant.

**Table 1.** Baseline demographics and disease characteristics.

|  | **Placebo**<br>**(N = 9)** | **E6742 100 mg**<br>**(N = 8)** | **E6742 200 mg**<br>**(N = 9)** | **Total**<br>**(N = 26)** |
| --- | --- | --- | --- | --- |
| Age (years), Mean (SD) | 38.9 (9.20) | 33.6 (12.97) | 40.4 (11.17) | 37.8 (11.07) |
| Female, n (%) | 8 (88.9) | 8 (100.0) | 8 (88.9) | 24 (92.3) |
| Ethnic origin (Asian), n (%) | 9 (100.0) | 8 (100.0) | 9 (100.0) | 26 (100.0) |
| Weight (kg), Mean (SD) | 53.6 (13.74) | 53.2 (6.34) | 51.8 (8.26) | 52.9 (9.70) |
| BMI (kg/m<sup>2</sup>), Mean (SD) | 21.1 (4.58) | 20.9 (2.55) | 21.1 (2.73) | 21.1 (3.31) |
| Disease Duration of SLE (years), Mean (SD) | 7.7 (6.10) | 4.5 (3.62) | 8.2 (6.96) | 6.9 (5.81) |
| SLEDAI-2K total score, Mean (SD) | 7.8 (2.91) | 8.6 (4.69) | 6.7 (2.83) | 7.7 (3.47) |
| BILAG-2004 components, n (%) |  |  |  |  |
| Mucocutaneous, A/B/C | 0/6/0 (66.7) | 0/6/1 (87.5) | 1/4/3 (88.9) | 1/16/4 (80.8) |
| Musculoskeletal, A/B/C | 0/4/3 (77.8) | 0/5/1 (75.0) | 0/4/1 (55.6) | 0/13/5 (69.2) |
| Haematological, A/B/C | 0/0/5 (55.6) | 0/0/5 (62.5) | 0/0/5 (55.6) | 0/0/15 (57.7) |
| PGA (0-3) score, Mean (SD) | 1.3 (0.50) | 1.0 (0.27) | 1.1 (0.40) | 1.2 (0.41) |
| CLASI activity score, Mean (SD) | 3.7 (4.69) | 3.5 (3.30) | 3.7 (4.77) | 3.6 (4.17) |
| Tender joint count (68), Mean (SD) | 9.6 (15.88) | 9.1 (12.38) | 5.4 (7.13) | 8.0 (11.98) |
| Swollen joint count (66), Mean (SD) | 2.9 (5.30) | 3.8 (4.98) | 1.8 (2.33) | 2.8 (4.28) |
| Autoantibody Titer, n (%) |  |  |  |  |
| ANA positive (≥ 80 Titer) | 7 (77.8) | 7 (87.5) | 7 (77.8) | 21 (80.8) |
| Anti-dsDNA IgG positive ($>$ 12 U/mL) | 2 (22.2) | 2 (25.0) | 3 (33.3) | 7 (26.9) |
| Anti-Sm positive (≥ 10 U/mL) | 3 (33.3) | 0 | 1 (11.1) | 4 (15.4) |
| Complement level, n (%) |  |  |  |  |
| Low C3 ($<$ 0.65 g/L) | 4 (44.4) | 3 (37.5) | 2 (22.2) | 9 (34.6) |
| Low C4 ($<$ 0.13 g/L) | 5 (55.6) | 4 (50.0) | 6 (66.7) | 15 (57.7) |
| Concomitant Medication for SLE, n (%) |  |  |  |  |
| Oral glucocorticoids, n (%) | 8 (88.9) | 8 (100.0) | 9 (100.0) | 25 (96.2) |
| Dose (mg/day)*, Mean (SD) | 4.8 (3.45) | 5.6 (2.15) | 6.2 (2.99) | 5.6 (2.85) |
| ≤10 mg/day, n (%) | 7 (77.8) | 8 (100.0) | 9 (100.0) | 24 (92.3) |
| $>$10 mg/day, n (%) | 1 (11.1) | 0 | 0 | 1 (3.8) |
| Hydroxychloroquine, n (%) | 8 (88.9) | 7 (87.5) | 6 (66.7) | 21 (80.8) |
| Immunosuppressants, n (%) | 4 (44.4) | 4 (50.0) | 4 (44.4) | 12 (46.2) |
| Mycophenolate mofetil | 0 | 0 | 1 (11.1) | 1 (3.8) |
| Methotrexate | 0 | 2 (25.0) | 1 (11.1) | 3 (11.5) |
| Tacrolimus | 4 (44.4) | 2 (25.0) | 2 (22.2) | 8 (30.8) |

Data expressed as n (%) or mean (standard deviation, SD).
\*Prednisolone or equivalent.
ANA, antinuclear antibody; anti-dsDNA, anti-double-stranded DNA; anti-Sm, anti-Smith; BILAG, British Isles Lupus Assessment Group; BMI, body mass index; CLASI, Cutaneous Lupus Erythematosus Disease Area and Severity Index; PGA, Physician's Global Assessment; SLE, systemic lupus erythematosus; SLEDAI, Systemic Lupus Erythematosus Disease Activity Index.

### Safety

Treatment-emergent adverse events (TEAEs) occurred in 6 of 9 patients (66.7%) in the placebo group, 3 of 8 patients (37.5%) in the 100 mg group, and 7 of 9 patients (77.8%) in the 200 mg group, with a similar incidence between the treatment groups (table 2). The TEAEs reported in 2 or more patients in either of the E6742 treatment groups, categorized as MedDRA Preferred Terms, were nasopharyngitis (25.0% [2/8 patients]) in the 100 mg group and headache (22.2% [2/9 patients]) in the 200 mg group. Although these TEAEs only occurred in E6742-treated patients, all but one (a case of nasopharyngitis in the 100 mg group) were considered unrelated to the study drug. All TEAEs were CTCAE Grade 1 or 2 in severity; no Grade 3 or higher TEAEs occurred. No deaths were reported during the study. One participant in the 200 mg group experienced pneumonia (serious AE, CTCAE Grade 2) that led to withdrawal of the study treatment, but recovered within 1 week after hospitalization. At each patient visit, 12-lead ECG assessments were performed. On Days 1 and 15, ECGs were measured at 3 hours post-dose, which roughly corresponded to the peak concentration (C_max_) of E6742. No patients in any treatment group had a clinically meaningful post-baseline corrected QT (QTc) interval using Fridericia’s formula exceeding 480 msec or an increase exceeding 60 msec (supplemental figure S1). No clinically significant findings in clinical laboratory tests, vital signs, or chest X-rays were observed in E6742-treated patients.

**Table 2.**
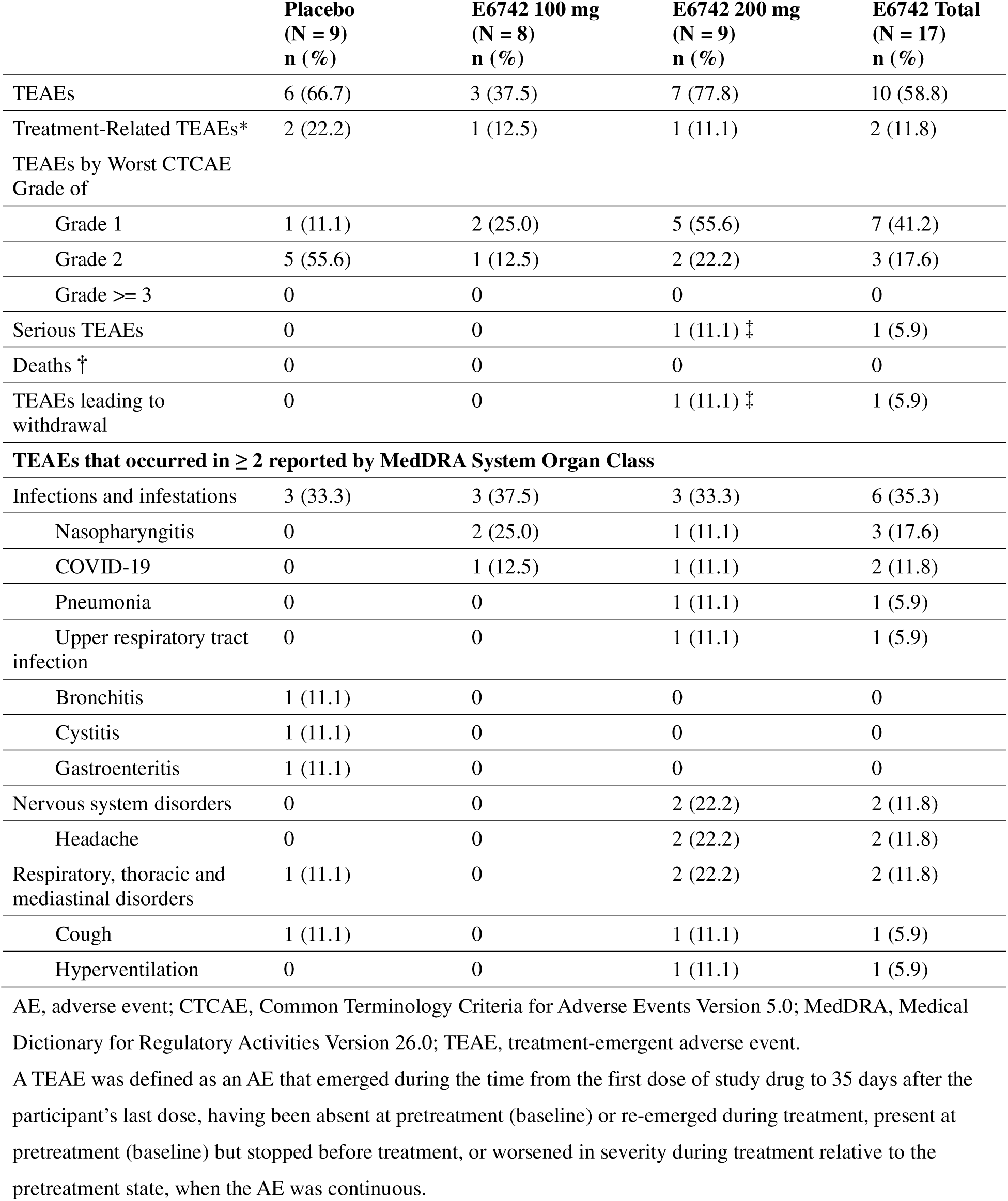
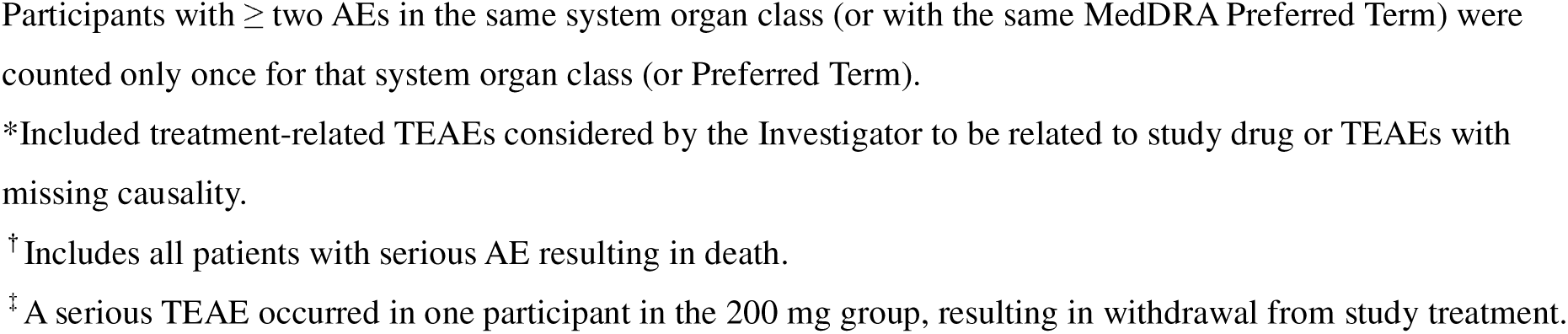
Summary of safety data.

|  | **Placebo**<br>**(N = 9)**<br>**n (%)** | **E6742 100 mg**<br>**(N = 8)**<br>**n (%)** | **E6742 200 mg**<br>**(N = 9)**<br>**n (%)** | **E6742 Total**<br>**(N = 17)**<br>**n (%)** |
| --- | --- | --- | --- | --- |
| TEAEs | 6 (66.7) | 3 (37.5) | 7 (77.8) | 10 (58.8) |
| Treatment-Related TEAEs* | 2 (22.2) | 1 (12.5) | 1 (11.1) | 2 (11.8) |
| TEAEs by Worst CTCAE Grade of |  |  |  |  |
| Grade 1 | 1 (11.1) | 2 (25.0) | 5 (55.6) | 7 (41.2) |
| Grade 2 | 5 (55.6) | 1 (12.5) | 2 (22.2) | 3 (17.6) |
| Grade ≥ 3 | 0 | 0 | 0 | 0 |
| Serious TEAEs | 0 | 0 | 1 (11.1) ‡ | 1 (5.9) |
| Deaths † | 0 | 0 | 0 | 0 |
| TEAEs leading to withdrawal | 0 | 0 | 1 (11.1) ‡ | 1 (5.9) |
| **TEAEs that occurred in ≥ 2 reported by MedDRA System Organ Class** |  |  |  |  |
| Infections and infestations | 3 (33.3) | 3 (37.5) | 3 (33.3) | 6 (35.3) |
| Nasopharyngitis | 0 | 2 (25.0) | 1 (11.1) | 3 (17.6) |
| COVID-19 | 0 | 1 (12.5) | 1 (11.1) | 2 (11.8) |
| Pneumonia | 0 | 0 | 1 (11.1) | 1 (5.9) |
| Upper respiratory tract infection | 0 | 0 | 1 (11.1) | 1 (5.9) |
| Bronchitis | 1 (11.1) | 0 | 0 | 0 |
| Cystitis | 1 (11.1) | 0 | 0 | 0 |
| Gastroenteritis | 1 (11.1) | 0 | 0 | 0 |
| Nervous system disorders | 0 | 0 | 2 (22.2) | 2 (11.8) |
| Headache | 0 | 0 | 2 (22.2) | 2 (11.8) |
| Respiratory, thoracic and mediastinal disorders | 1 (11.1) | 0 | 2 (22.2) | 2 (11.8) |
| Cough | 1 (11.1) | 0 | 1 (11.1) | 1 (5.9) |
| Hyperventilation | 0 | 0 | 1 (11.1) | 1 (5.9) |

AE, adverse event; CTCAE, Common Terminology Criteria for Adverse Events Version 5.0; MedDRA, Medical Dictionary for Regulatory Activities Version 26.0; TEAE, treatment-emergent adverse event.
A TEAE was defined as an AE that emerged during the time from the first dose of study drug to 35 days after the participant's last dose, having been absent at pretreatment (baseline) or re-emerged during treatment, present at pretreatment (baseline) but stopped before treatment, or worsened in severity during treatment relative to the pretreatment state, when the AE was continuous.
Participants with ≥ two AEs in the same system organ class (or with the same MedDRA Preferred Term) were counted only once for that system organ class (or Preferred Term).
\*Included treatment-related TEAEs considered by the Investigator to be related to study drug or TEAEs with missing causality.
<sup>†</sup> Includes all patients with serious AE resulting in death.
<sup>‡</sup> A serious TEAE occurred in one participant in the 200 mg group, resulting in withdrawal from study treatment.

### PK

The median time for E6742 to reach C_max_ (t_max_) and median time to steady-state maximum concentration (t_ss,max_) occurred at 1.44–1.87 hours and 1.11–1.42 hours, respectively, following oral dosing at 100 and 200 mg BID. Plasma concentrations of E6742 increased with increasing doses (supplemental figure S2). The geometric mean accumulation ratio of C_max_ and area under the curve of E6742 were 1.03–1.32 and 1.04–1.28, respectively (data not shown).

### Biomarker response

The 127 SLE-specific IGSs were evaluated in this study. An early decrease in the expression of the majority these genes was observed from 2 weeks post-treatment onwards, with the reductions maintained throughout the treatment with E6742. For example, the expression levels of *MX1*, *IFIT1*, and *IFI44L* were markedly reduced at Week 12 in the E6742-treated groups, showing the following median percent changes from baseline: –87.6% in the 100 mg group, –86.1% in the 200 mg group, and 6.83% in the placebo group for *MX1*; –88.5% in the 100 mg group, –90.7% in the 200 mg group, and 14.8% in the placebo group for *IFIT1*; and –95.5% in the 100 mg group, –95.9% in the 200 mg group, and 14.8% in the placebo group for *IFI44L* (data not shown). Furthermore, IGS scores based on the major 21 genes immediately dropped under treatment with E6742 (figure 2), with both the 100 mg and 200 mg groups showing approximately 90% reductions by 2 weeks post-treatment; these reductions were maintained throughout the treatment period.

**Figure 2.**
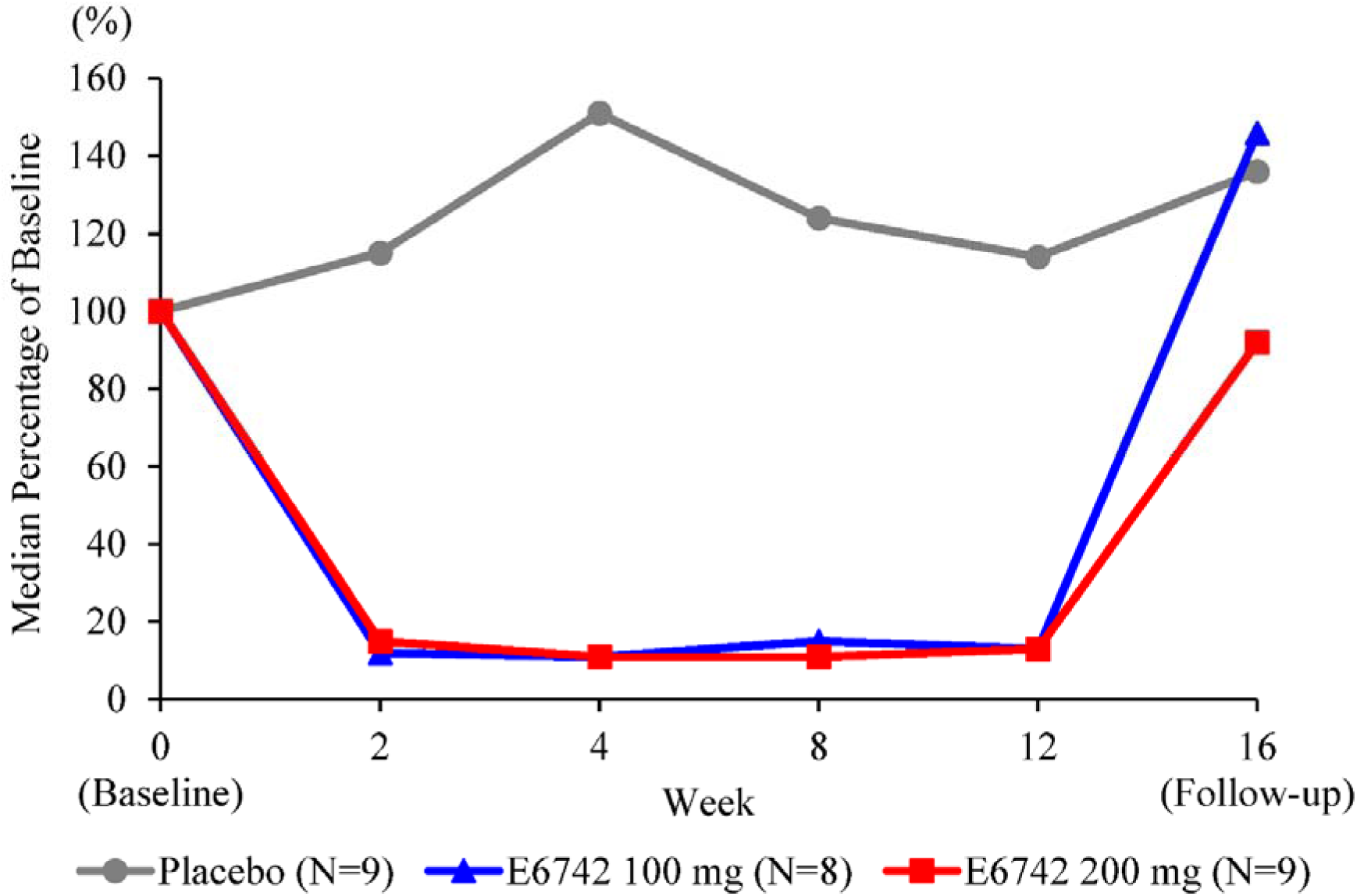
Effect of E6742 on IGS scores in patients with SLE. The interferon gene signature (IGS) scores were calculated from the fold change in expression of a set of 21 IGSs. Only patients with non-missing data at both baseline and the relevant post-baseline visits were included in the calculation. Results were expressed as the median percent change from baseline. SLE, systemic lupus erythematosus. The actual data of each plot are included in supplemental table S2.

Pharmacodynamic biomarkers were also assessed using an ex-vivo assay. R848, a TLR7/8 agonist, was incubated with blood samples collected on Days 1 and 15 to evaluate the inhibitory effect of E6742 on the levels of R848-induced cytokines (IL-1β, IL-6, and TNF-α). The mean concentrations of IL-1β, IL-6, and TNF-α were significantly decreased from baseline in E6742-treated participants at 1 hour post-dose on Day 1, with mean percent changes of > 95%. These reductions were generally maintained until Day 15 (figure 3).

**Figure 3.**
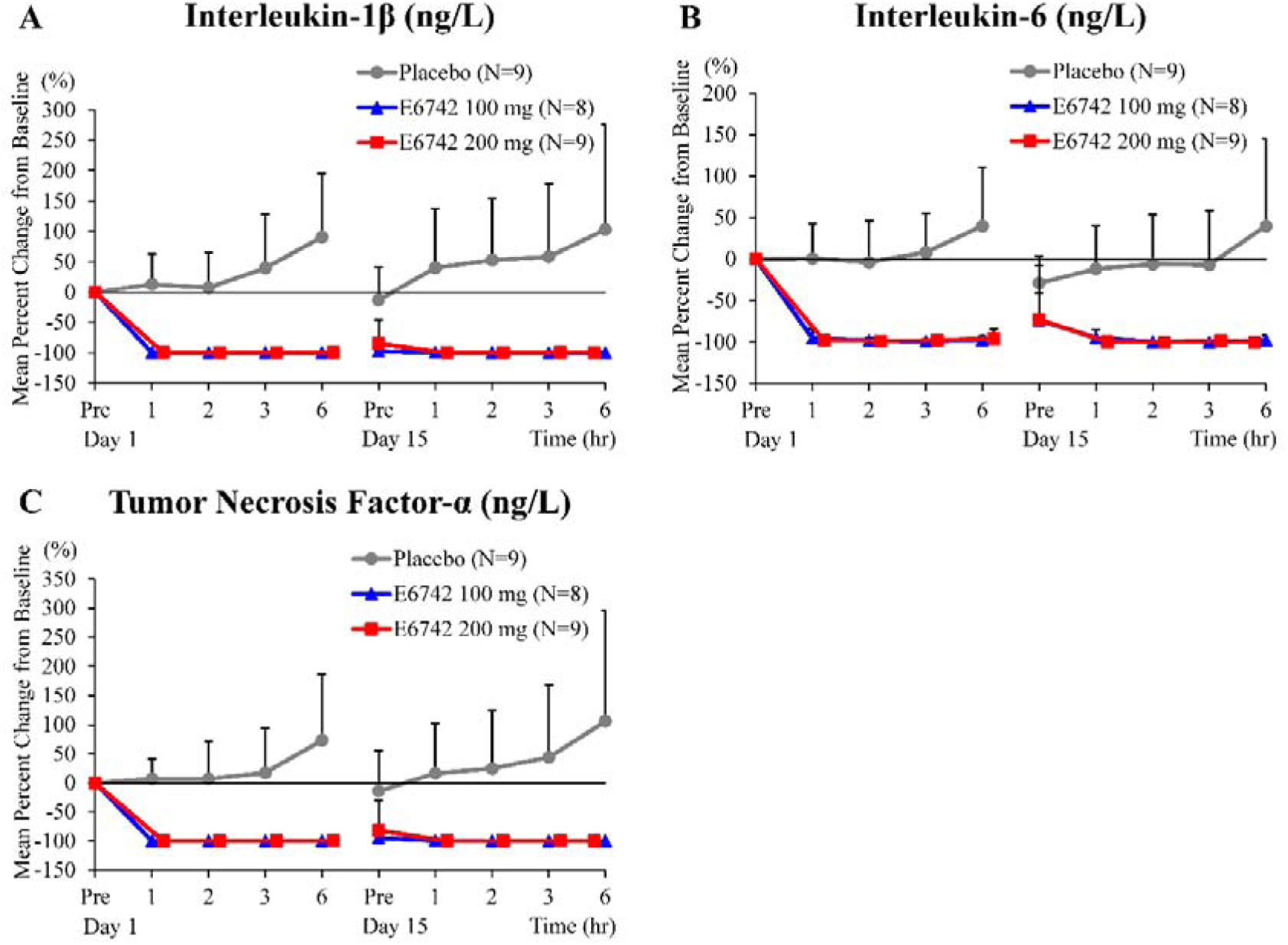
Effects of E6742 on R848-mediated ex-vivo induction of blood cytokines: (A) interleukin-1β, (B) interleukin-6, and (C) tumor necrosis factor-α. Linear plots of mean (+ standard deviation) percent change from baseline of the concentrations of the indicated cytokines following treatment of Day 1 and Day 15 blood samples with Toll-like receptor 7/8 agonist R848. All descriptive statistics were calculated using the percent change from baseline of the median measured concentration in the analytical sample (n = 3). If 1/3 measurements were below the limit of quantification (BLQ), the BLQ was assigned as zero and the median value was used; if 2/3 were BLQ, the lower limit of quantification was used; if all measurements were BLQ, zero was used. Pre, pre-dose. The actual data of each plot are included in supplemental table S3.

### Efficacy

At Week 12, a BICLA response was observed in 33.3% (3/9) of patients in the placebo group, 37.5% (3/8) in the 100 mg group, and 57.1% (4/7) in the 200 mg group, with higher rates in both E6742 treatment groups compared with that in the placebo group (figure 4). There was also a dose–response trend between BICLA response and E6742 dose level at Week 12. Two patients in the 200 mg group who had no A or B category BILAG-2004 scores at baseline were excluded from the BICLA response calculation.

**Figure 4.**
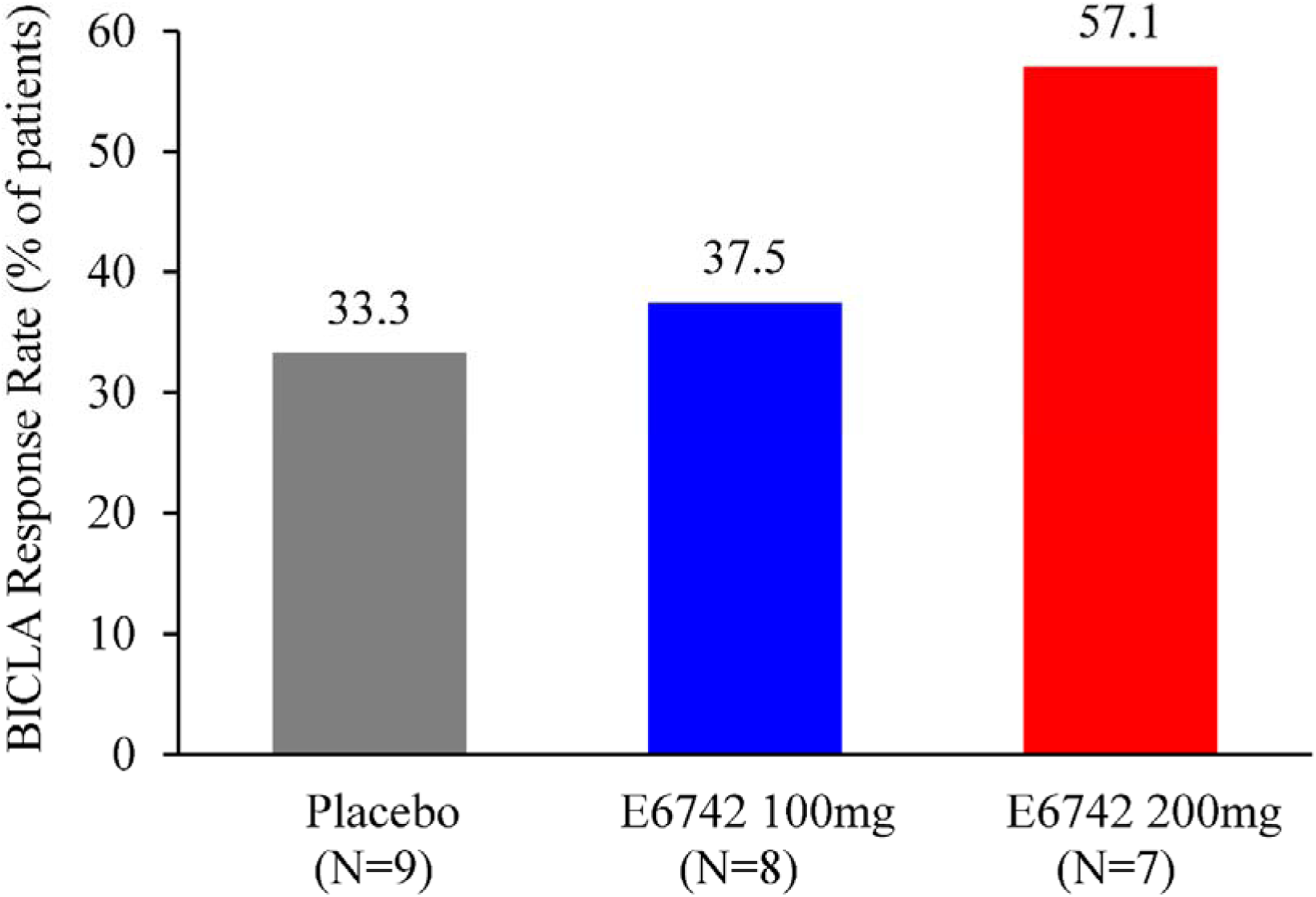
BICLA response at Week 12 of treatment with E6742. Response rate analyzed for patients with at least 1 A or B category score in the British Isles Lupus Assessment Group Index 2004 (BILAG-2004) at baseline. Participants who discontinued treatment were imputed as non-responders for all of the visits following treatment discontinuation. BICLA, British Isles Lupus Assessment Group-based Composite Lupus Assessment.

Clinical outcomes, including PGA scores, CLASI activity scores, and joint counts, generally improved from baseline in E6742-treated patients compared with those in placebo-treated patients (table 3). Additionally, serological parameters including anti-dsDNA antibodies and complement C3 and C4 levels showed improvements in the E6742-treated group. At Week 12, the reduction from baseline in CLASI activity score in the 200 mg group (–2.4) was larger than that in the placebo group (–1.3). The CLASI-50 response rates were higher in the E6742 100 mg (57.1%) and 200 mg (50.0%) groups than in the placebo group (33.3%). The reductions from baseline in tender joint counts in the 100 mg and 200 mg groups (–2.4 and –5.3, respectively) were larger compared with that in the placebo group (–0.9) at Week 12. Regarding swollen joint counts, both E6742 treatment groups exhibited large reductions (–3.8 at 100 mg and –1.5 at 200 mg), whereas the placebo group showed a slight increase (0.1) at Week 12. In both E6742-treated patients who had swollen joint counts at baseline, all joint swelling had disappeared at Week 12. Clear reductions in anti-dsDNA immunoglobulin G antibodies were also observed in the E6742 treated groups (–7.7 at 100 mg and –2.7 at 200 mg) at Week 12.

**Table 3.** Clinical outcomes at Week 12.

| **Outcomes** | **Placebo**<br>**(N=9)** | **E6742 100 mg**<br>**(N=8)** | **E6742 200 mg**<br>**(N=9)\*** |
| --- | --- | --- | --- |
| Change from baseline at Week 12 †, Mean (SD) |  |  |  |
| SLEDAI-2K total score | -2.2 (2.11) | -1.6 (2.26) | -2.3 (2.25) |
| PGA score | -0.4 (0.45) | -0.6 (0.45) | -0.7 (0.38) |
| CLASI activity score | -1.3 (2.18) | -1.1 (1.13) | -2.4 (4.78) |
| Tender joint counts (out of 68 joints) | -0.9 (9.97) | -2.4 (7.84) | -5.3 (7.17) |
| Swollen joint counts (out of 66 joints) | 0.1 (1.27) | -3.8 (4.98) | -1.5 (2.33) |
| Anti-dsDNA IgG antibody (IU/mL) | 2.3 (6.78) | -7.7 (12.89) | -2.7 (5.35) |
| Complement C3 (g/L) | 0.02 (0.16) | 0.06 (0.072) | 0.06 (0.16) |
| Complement C4 (g/L) | -0.008 (0.032) | 0.009 (0.024) | 0.004 (0.038) |
| Responder rate at Week 12, n/m (%) |  |  |  |
| CLASI-50 response ‡ | 2/6 (33.3) | 4/7 (57.1) | 4/8 (50.0) |

\*Includes one participant with missing data due to early discontinuation.
† Only patients with non-missing data at both baseline and the relevant post-baseline visit were included in the change from baseline summary statistics.
‡ Response rate analyzed a decrease of $\geq 50\%$ from baseline Cutaneous Lupus Erythematosus Disease Area and Severity Index (CLASI) activity score for patients with $\geq 1$ point in CLASI anti-dsDNA, anti-double-stranded DNA; PGA, Physician's Global Assessment; SLEDAI-2K, Systemic Lupus Erythematosus Disease Activity Index 2000.

## DISCUSSION

E6742 is a small molecule compound that directly binds to the single-stranded RNA-sensing receptors TLR7 and TLR8, inhibiting activation of the respective signaling pathways. This multicenter, randomized, double-blind, placebo-controlled study provides the first clinical evidence of E6742 in the treatment of SLE.

E6742 was generally favorable and well tolerated throughout the 12-week treatment period. There was no clear difference in the incidence of TEAEs among the 100 mg, 200 mg, and placebo groups (table 2). A nonfatal but serious TEAE of pneumonia (CTCAE Grade 2) occurred in one participant in the 200 mg group, who recovered within about 1 week after hospitalization. Because the risk of infection is a major problem with currently available SLE treatments, namely glucocorticoids, immunosuppressants, and biologics, the possible risk of infection with E6742 should be noted. Although inhibition of TLRs, as key sensors of viruses and bacteria, might be expected to increase infection risk, selective TLR inhibition might decrease the potential infection risks. Furthermore, there is redundancy in the immune system that ensures protection against infections. Thus, compared to existing nonspecific immunosuppressive therapies or biologics, which target specific cytokines, E6742 may offer a reduced risk of infection through its selective inhibition of TLR7/8, which act upstream in the immune cascade and exhibit hyperactivity in SLE. Because the case of pneumonia observed in this trial was bacterial in nature, the possible relationship between this pneumonia and inhibition of TLR7/8, as a sensor of virus, is inconclusive. The overall incidences of infection were generally comparable in the placebo (33.3%) and E6742 total (35.3%) groups (table 2), and there were no changes in laboratory values related to infection risk, including neutrophil and white blood cell counts, after E6742 administration.

Because multiple drugs are often used concomitantly in the treatment of SLE, it was important to evaluate the safety of E6742 when used in combination with other drugs. Considering the results of our previous phase 1 studies of E6742, the non-linear PK at high doses and the serum concentration-dependent risk of QT prolongation of E6742 were flagged as potential safety concerns [19]. According to the US Food and Drug Administration, HCQ can cause abnormal heart rhythms such as QT interval prolongation and a dangerously rapid heart rate called ventricular tachycardia [24]. In this trial, 87.5% of patients in the 100 mg group and 66.7% of patients in the 200 mg group concomitantly used HCQ. However, there were no specific concerns regarding PK parameters after multiple oral doses of E6742 in SLE patients, which were similar to those in healthy adults. Likewise, there were no QTc safety signals after E6742 treatment. These results suggested that E6742 may be indicated as a viable therapeutic agent in real-world clinical practice. However, there are three main limitations regarding the interpretations of E6742 safety data from this trial: 1) only Japanese patients were enrolled; 2) patients with specific complications, such as severe lupus nephritis and central nervous system lupus, were excluded; and 3) the sample size and evaluation period were limited. Therefore, a longer and more detailed evaluation is required to fully derive the safety profile of E6742.

The heterogeneity of disease expression in SLE has made it challenging to find treatments with clear clinical efficacy. Therefore, as a secondary endpoint of this study we evaluated changes in biomarkers in response to E6742 treatment. Activation of the type I IFN system plays an important role in the immune system and is associated with the pathogenesis of SLE [25]. Specifically, patients with high IFN signaling are more likely to exhibit a disease flare of SLE [26] and IFN pathway activation has been shown to impart resistance to glucocorticoids [18, 25, 27]. TLR7 is highly expressed in pDCs and is thought to be a major trigger for type I IFN production in SLE [26], which is quantified using the IGS [28]. In this trial, IGS were immediately downregulated under E6742 treatment. Furthermore, although a direct comparison is not appropriate, the magnitude of change in IGS score from baseline (figure 2) was comparable to that reported for anti-IFN-α receptor antibodies, which directly suppress IFN signaling [29]. In addition, the concentrations of proinflammatory cytokines (IL-1β, IL-6, and TNF-α) were substantially (> 95%) reduced in the ex-vivo assay of TLR7/8 agonist challenge. With regard to clinical efficacy parameters, E6742 showed a dose-dependent improvement in the proportion of patients with a BICLA response at Week 12, with 33.3% in the placebo group, 37.5% in the E6742 100 mg group, and 57.1% in the E6742 200 mg group (figure 4). Clinical improvements in skin inflammation (CLASI activity score), arthritis (tender and swollen joints), and serological indicators, such as anti-double-stranded DNA antibody and complement levels, were also observed (table 3). These beneficial effects of E6742 in SLE suggest that E6742 indirectly regulates T-cell activation and B-cell differentiation by regulating the expression of IFN-related genes and proinflammatory cytokines [5].

Despite some limitations, including the short-term evaluation of efficacy, lack of disease activity score specified as an inclusion criterion, and the lack of stratification by baseline disease activity, the efficacy signals obtained in this trial were encouraging. Furthermore, the significant change in IGS in response to E6742 treatment warrants the aggressive evaluation of E6742 efficacy in additional patient populations with more active disease and diverse clinical phenotypes, which may substantiate its potential to improve disease flares and reduce glucocorticoids.

In conclusion, this phase 1/2 trial evaluated the safety, tolerability, PK, biomarker response, and efficacy profile of E6742 in patients with SLE, achieving the primary endpoint. These findings support further investigations of E6742 in the treatment of SLE.

## ACKNOWLEDGMENTS

The authors thank all of the investigators involved in this clinical trial of E6742 (supplemental list of investigators and study sites).

## DATA AVAILABILITY

Data are available upon reasonable request. The datasets used and/or analyzed during the current study are available from the corresponding author and sponsor on reasonable request.

## REFERENCES

1. Barber MRW, Drenkard C, Falasinnu T, et al. Global epidemiology of systemic lupus erythematosus. Nat Rev Rheumatol 2021;17(9):515–32.

2. Carter EE, Barr SG, Clarke AE. The global burden of SLE: prevalence, health disparities and socioeconomic impact. Nat Rev Rheumatol 2016;12(10):605–20.

3. Tanaka Y, Mizukami A, Kobayashi A, et al. Disease severity and economic burden in Japanese patients with systemic lupus erythematosus: a retrospective, observational study. Int J Rheum Dis 2018;21(8):1609–18.

4. Fanouriakis A, Kostopoulou M, Andersen J, et al. EULAR recommendations for the management of systemic lupus erythematosus: 2023 update. Ann Rheum Dis 2024;83(1):15–29.

5. Tanaka Y. State-of-the-art treatment of systemic lupus erythematosus. Int J Rheum Dis 2020;23(4):465–71.

6. Tanaka Y, O’Neill S, Li M, et al. Systemic lupus erythematosus: targeted literature review of the epidemiology, current treatment, and disease burden in the Asia Pacific region. Arthritis Care Res (Hoboken) 2022;74(2):187–98.

7. Mok CC, Teng YKO, Saxena R, et al. Treatment of lupus nephritis: consensus, evidence and perspectives. Nat Rev Rheumatol 2023;19(4):227–38.

8. Lund J, Sato A, Akira S, et al. Toll-like receptor 9-mediated recognition of herpes simplex virus-2 by plasmacytoid dendritic cells. J Exp Med 2003;198(3):513–20.

9. Heil F, Hemmi H, Hochrein H, et al. Species-specific recognition of single-stranded RNA via toll-like receptor 7 and 8. Science 2004;303(5663):1526–29.

10. El-Zayat SR, Sibaii H, Mannaa FA. Toll-like receptors activation, signaling, and targeting: an overview. Bull Natl Res Cent 2019;43(1):187.

11. Barrat FJ, Meeker T, Gregorio J, et al. Nucleic acids of mammalian origin can act as endogenous ligands for toll-like receptors and may promote systemic lupus erythematosus. J Exp Med 2005;202(8):1131–9.

12. Sakata K, Nakayamada S, Miyazaki Y, et al. Up-regulation of TLR7-mediated IFN-alpha production by plasmacytoid dendritic cells in patients with systemic lupus erythematosus. Front Immunol 2018;9:1957.

13. Ronnblom L, Leonard D. Interferon pathway in SLE: one key to unlocking the mystery of the disease. Lupus Sci Med 2019;6(1):e000270.

14. Postal M, Vivaldo JF, Fernandez-Ruiz R, et al. Type I interferon in the pathogenesis of systemic lupus erythematosus. Curr Opin Immunol 2020;67:87–94.

15. Shen N, Fu Q, Deng Y, et al. Sex-specific association of X-linked toll-like receptor 7 (TLR7) with male systemic lupus erythematosus. Proc Natl Acad Sci USA 2010;107(36):15838–43.

16. Brown GJ, Canete PF, Wang H, et al. TLR7 gain-of-function genetic variation causes human lupus. Nature 2022;605(7909):349–56.

17. Bender AT, Tzvetkov E, Pereira A, et al. TLR7 and TLR8 differentially activate the IRF and NF-kappaB pathways in specific cell types to promote inflammation. Immunohorizons 2020;4(2):93–107.

18. Deshmukh A, Pereira A, Geraci N, et al. Preclinical evidence for the glucocorticoid-sparing potential of a dual toll-like receptor 7/8 inhibitor in autoimmune diseases. J Pharmacol Exp Ther 2024;388:751–64.

19. Yamakawa N, Tago F, Nakai K, et al. First-in-human study of the safety, tolerability, pharmacokinetics, and pharmacodynamics of E6742, a dual antagonist of toll-like receptors 7 and 8, in healthy volunteers. Clin Pharmacol Drug Dev 2023;12(4):363–75.

20. Yao Y, Higgs BW, Morehouse C, et al. Development of potential pharmacodynamic and diagnostic markers for anti-IFN-alpha monoclonal antibody trials in systemic lupus erythematosus. Hum Genomics Proteomics 2009;2009:374312.

21. Furie R, Khamashta M, Merrill JT, et al. Anifrolumab, an anti-interferon-alpha receptor monoclonal antibody, in moderate-to-severe systemic lupus erythematosus. Arthritis Rheumatol 2017;69(2):376–86.

22. Kennedy WP, Maciuca R, Wolslegel K, et al. Association of the interferon signature metric with serological disease manifestations but not global activity scores in multiple cohorts of patients with SLE. Lupus Sci Med 2015;2(1):e000080.

23. Yao Y, Higgs BW, Richman L, et al. Use of type I interferon-inducible mRNAs as pharmacodynamic markers and potential diagnostic markers in trials with sifalimumab, an anti-IFNα antibody, in systemic lupus erythematosus. Arthritis Res Ther 2010;12 Suppl 1(Suppl 1):S6.

24. Food and Drug Administration: FDA cautions against use of hydroxychloroquine or chloroquine for COVID-19 outside of the hospital setting or a clinical trial due to risk of heart rhythm problems [online]. 2020. https://www.fda.gov/drugs/drug-safety-and-availability/fda-cautions-against-use-hydroxychloroquine-or-chloroquine-covid-19-outside-hospital-setting-or (accessed 29 Mar 2024).

25. Tanaka Y, Kusuda M, Yamaguchi Y. Interferons and systemic lupus erythematosus: Pathogenesis, clinical features, and treatments in interferon-driven disease. Mod Rheumatol 2023;33(5):857–67.

26. Landolt-Marticorena C, Bonventi G, Lubovich A, et al. Lack of association between the interferon-alpha signature and longitudinal changes in disease activity in systemic lupus erythematosus. Ann Rheum Dis 2009;68(9):1440–6.

27. Dankers W, Northcott M, Bennett T, et al. Type 1 interferon suppresses expression and glucocorticoid induction of glucocorticoid-induced leucine zipper (GILZ). Front Immunol 2022;13:1034880.

28. Vital EM, Merrill JT, Morand EF, et al. Anifrolumab efficacy and safety by type I interferon gene signature and clinical subgroups in patients with SLE: post hoc analysis of pooled data from two phase III trials. Ann Rheum Dis 2022;81(7):951–61.

29. Morand EF, Furie R, Tanaka Y, et al. Trial of anifrolumab in active systemic lupus erythematosus. N Engl J Med 2020;382(3):211–21.

