## supplemental for "Safety, Pharmacokinetics, Biomarker Response, and Efficacy of E6742, a Dual Antagonist of Toll-Like Receptors 7 and 8, in a First-in-Patient, Randomized, Double-Blind, Phase 1/2 Study in Systemic Lupus Erythematosus"

### SUPPLEMENTAL MATERIAL

#### List of investigators and study sites

| List of Investigators | Site Number | Facility |
|---|---|---|
| Shingo, Nakayamada | 1001 | Hospital of the University of Occupational and Environmental Health, Japan |
| Masanari, Kodera | 1002 | Japan Community Health Care Organization Chukyo Hospital |
| Yoichiro, Haji | 1003 | Daido Clinic |
| Kensuke, Oryoji (20May2022-25May2023) | 1004 | Matsuyama Red Cross Hospital |
| Shinichi, Mizuki (26May2023-) | | |
| Masaru Kato | 1005 | Hokkaido University Hospital |
| Tomonori, Ishii | 1006 | Tohoku University Hospital |
| Naoto, Yokogawa | 1007 | Tokyo Metropolitan Tama Medical Center |
| Tomoya, Miyamura | 1008 | National Hospital Organization Kyushu Medical Center |
| Yasuhiro, Kato | 1009 | Osaka University Hospital |
| Futoshi, Iwata | 1010 | St. Luke’s International Hospital |
| Hiroshi, Kaneko | 1011 | National Center for Global Health and Medicine |
| Kentaro Minowa | 1012 | Juntendo University Hospital |

**Supplemental Table S1 List of interferon gene signature evaluated in this study**

| Ensembl Gene ID | Gene Symbol |
|---|---|
| ENSG00000160710 | ADAR |
| ENSG00000163568 | AIM2 |
| ENSG00000152766 | ANKRD22 |
| ENSG00000128383 | APOBEC3A |
| ENSG00000100342 | APOL1 |
| ENSG00000140750 | ARHGAP17 |
| ENSG00000168062 | BATF2 |
| ENSG00000198604 | BAZ1A |
| ENSG00000282851 | BISPR |
| ENSG00000106605 | BLVRA |
| ENSG00000130303 | BST2 |
| ENSG00000197536 | C5orf56 |
| ENSG00000163823 | CCR1 |
| ENSG00000086065 | CHMP5 |
| ENSG00000134326 | CMPK2 |
| ENSG00000137200 | CMTR1 |
| ENSG00000173198 | CYSLTR1 |
| ENSG00000107201 | DDX58 |
| ENSG00000137628 | DDX60 |
| ENSG00000181381 | DDX60L |
| ENSG00000108771 | DHX58 |
| ENSG00000175550 | DRAP1 |
| ENSG00000163840 | DTX3L |
| ENSG00000055332 | EIF2AK2 |
| ENSG00000133106 | EPSTI1 |
| ENSG00000010030 | ETV7 |
| ENSG00000116663 | FBXO6 |
| ENSG00000198019 | FCGR1B |
| ENSG00000117228 | GBP1 |
| ENSG00000130589 | HELZ2 |
| ENSG00000138646 | HERC5 |
| ENSG00000138642 | HERC6 |
| ENSG00000163666 | HESX1 |

| Ensembl Gene ID | Gene Symbol |
|---|---|
| ENSG00000196684 | HSH2D |
| ENSG00000163565 | IFI16 |
| ENSG00000165949 | IFI27 |
| ENSG00000068079 | IFI35 |
| ENSG00000137965 | IFI44 |
| ENSG00000137959 | IFI44L |
| ENSG00000126709 | IFI6 |
| ENSG00000115267 | IFIH1 |
| ENSG00000185745 | IFIT1 |
| ENSG00000119922 | IFIT2 |
| ENSG00000119917 | IFIT3 |
| ENSG00000152778 | IFIT5 |
| ENSG00000185885 | IFITM1 |
| ENSG00000185201 | IFITM2 |
| ENSG00000142089 | IFITM3 |
| ENSG00000136689 | IL1RN |
| ENSG00000185507 | IRF7 |
| ENSG00000213928 | IRF9 |
| ENSG00000187608 | ISG15 |
| ENSG00000172183 | ISG20 |
| ENSG00000165185 | KIAA1958 |
| ENSG00000127528 | KLF2 |
| ENSG00000130487 | KLHDC7B |
| ENSG00000117009 | KMO |
| ENSG00000078081 | LAMP3 |
| ENSG00000002549 | LAP3 |
| ENSG00000108679 | LGALS3BP |
| ENSG00000168961 | LGALS9 |
| ENSG00000205837 | LINC00487 |
| ENSG00000160932 | LY6E |
| ENSG00000140280 | LYSMD2 |
| ENSG00000157601 | MX1 |
| ENSG00000183486 | MX2 |

| Ensembl Gene ID | Gene Symbol |
|---|---|
| ENSG00000102921 | N4BP1 |
| ENSG00000124357 | NAGK |
| ENSG00000111912 | NCOA7 |
| ENSG00000123609 | NMI |
| ENSG00000089127 | OAS1 |
| ENSG00000111335 | OAS2 |
| ENSG00000111331 | OAS3 |
| ENSG00000135114 | OASL |
| ENSG00000177989 | ODF3B |
| ENSG00000115155 | OTOF |
| ENSG00000178685 | PARP10 |
| ENSG00000059378 | PARP12 |
| ENSG00000173193 | PARP14 |
| ENSG00000138496 | PARP9 |
| ENSG00000136147 | PHF11 |
| ENSG00000145287 | PLAC8 |
| ENSG00000188313 | PLSCR1 |
| ENSG00000140464 | PML |
| ENSG00000138035 | PNPT1 |
| ENSG00000105287 | PRKD2 |
| ENSG00000240065 | PSMB9 |
| ENSG00000092010 | PSME1 |
| ENSG00000100911 | PSME2 |
| ENSG00000266094 | RASSF5 |
| ENSG00000125826 | RBCK1 |
| ENSG00000143344 | RGL1 |
| ENSG00000134321 | RSAD2 |
| ENSG00000136514 | RTP4 |
| ENSG00000205413 | SAMD9 |
| ENSG00000177409 | SAMD9L |
| ENSG00000284194 | SCO2 |
| ENSG00000149131 | SERPING1 |
| ENSG00000130813 | SHFL |

| Ensembl Gene ID | Gene Symbol |
|---|---|
| ENSG00000164054 | SHISA5 |
| ENSG00000088827 | SIGLEC1 |
| ENSG00000135899 | SP110 |
| ENSG00000079263 | SP140 |
| ENSG00000196141 | SPATS2L |
| ENSG00000115415 | STAT1 |
| ENSG00000170581 | STAT2 |
| ENSG00000168394 | TAP1 |
| ENSG00000196116 | TDRD7 |
| ENSG00000196664 | TLR7 |
| ENSG00000121858 | TNFSF10 |
| ENSG00000102524 | TNFSF13B |
| ENSG00000136816 | TOR1B |
| ENSG00000135148 | TRAFD1 |
| ENSG00000168016 | TRANK1 |
| ENSG00000132109 | TRIM21 |
| ENSG00000132274 | TRIM22 |
| ENSG00000121236 | TRIM6 |
| ENSG00000185880 | TRIM69 |
| ENSG00000025708 | TYMP |
| ENSG00000156587 | UBE2L6 |
| ENSG00000184979 | USP18 |
| ENSG00000028116 | VRK2 |
| ENSG00000132530 | XAF1 |
| ENSG00000124256 | ZBP1 |
| ENSG00000105939 | ZC3HAV1 |
| ENSG00000141664 | ZCCHC2 |
| ENSG00000124201 | ZNFX1 |

**Supplemental Table S2 Median percent change from baseline for IGS score**

| Visit | Placebo (N=9) | E6742 100 mg BID (N=8) | E6742 200 mg BID (N=9) |
|---|---|---|---|
| Baseline | | | |
| n | 9 | 8 | 9 |
| Median Percentage | 100 | 100 | 100 |
| Week 2 | | | |
| n | 9 | 8 | 9 |
| Median Percentage | 115 | 12 | 15 |
| Week 4 | | | |
| n | 9 | 8 | 9 |
| Median Percentage | 151 | 11 | 11 |
| Week 8 | | | |
| n | 9 | 8 | 9 |
| Median Percentage | 124 | 15 | 11 |
| Week 12 | | | |
| n | 9 | 8 | 8 |
| Median Percentage | 114 | 13 | 13 |
| Follow-up | | | |
| n | 9 | 8 | 9 |
| Median Percentage | 136 | 146 | 92 |

**Supplemental Table S3 Mean percent change from baseline for R848-mediated ex-vivo induction of blood cytokines**

| Visit / Timepoint<br>Statistics | Placebo<br>(N=9) | E6742 100 mg BID<br>(N=8) | E6742 200 mg BID<br>(N=9) |
|---|---|---|---|
| **Interleukin-1β (ng/L)** | | | |
| Day 1 / pre-dose (Baseline) | | | |
| n | 9 | 8 | 9 |
| Mean (SD) | 0 | 0 | 0 |
| Day 1 / 1 hr post-dose | | | |
| n | 9 | 8 | 9 |
| Mean (SD) | 12.872 (50.3489) | -99.530 (1.0091) | -99.471 (0.8017) |
| Day 1 / 2 hrs post-dose | | | |
| n | 9 | 8 | 9 |
| Mean (SD) | 7.409 (58.1975) | -99.704 (0.5501) | -99.757 (0.3921) |
| Day 1 / 3 hrs post-dose | | | |
| n | 9 | 8 | 9 |
| Mean (SD) | 39.749 (88.3659) | -99.449 (1.0818) | -99.669 (0.6047) |
| Day 1 / 6 hrs post-dose | | | |
| n | 9 | 8 | 9 |
| Mean (SD) | 90.754 (103.6237) | -99.229 (1.5020) | -99.500 (1.5000) |
| Day 15 / pre-dose | | | |
| n | 9 | 8 | 9 |
| Mean (SD) | -12.961 (54.2801) | -97.337 (2.0642) | -85.065 (39.8342) |
| Day 15 / 1 hr post-dose | | | |
| n | 9 | 8 | 9 |
| Mean (SD) | 40.400 (96.9953) | -99.113 (1.3800) | -99.804 (0.2574) |
| Day 15 / 2 hrs post-dose | | | |
| n | 9 | 8 | 9 |
| Mean (SD) | 52.855 (100.7442) | -99.539 (0.5360) | -99.827 (0.3185) |
| Day 15 / 3 hrs post-dose | | | |
| n | 9 | 8 | 9 |
| Mean (SD) | 58.409 (119.9955) | -99.884 (0.3274) | -99.353 (1.2534) |
| Day 15 / 6 hrs post-dose | | | |
| n | 9 | 8 | 9 |
| Mean (SD) | 102.605 (172.5928) | -99.722 (0.5472) | -99.609 (0.8670) |
| **Interleukin-6 (ng/L)** | | | |
| Day 1 / pre-dose (Baseline) | | | |
| n | 9 | 8 | 9 |
| Mean (SD) | 0 | 0 | 0 |
| Day 1 / 1 hr post-dose | | | |
| n | 9 | 8 | 9 |
| Mean (SD) | 1.014 (42.0451) | -95.615 (11.6667) | -97.877 (5.5573) |

| Visit / Timepoint<br>Statistics | Placebo<br>(N=9) | E6742 100 mg BID<br>(N=8) | E6742 200 mg BID<br>(N=9) |
|---|---|---|---|
| Day 1 / 2 hrs post-dose | | | |
| n | 9 | 8 | 9 |
| Mean (SD) | -3.625 (50.1343) | -98.216 (3.3470) | -98.812 (2.0384) |
| Day 1 / 3 hrs post-dose | | | |
| n | 9 | 8 | 9 |
| Mean (SD) | 8.364 (47.0364) | -99.312 (0.9373) | -98.008 (4.9924) |
| Day 1 / 6 hrs post-dose | | | |
| n | 9 | 8 | 9 |
| Mean (SD) | 40.270 (70.4681) | -97.444 (4.7430) | -95.809 (11.6223) |
| Day 15 / pre-dose | | | |
| n | 9 | 8 | 9 |
| Mean (SD) | -28.732 (32.4104) | -73.919 (33.1609) | -73.110 (65.7286) |
| Day 15 / 1 hr post-dose | | | |
| n | 9 | 8 | 9 |
| Mean (SD) | -11.394 (51.1945) | -94.890 (9.5397) | -99.787 (0.2984) |
| Day 15 / 2 hrs post-dose | | | |
| n | 9 | 8 | 9 |
| Mean (SD) | -5.813 (59.5118) | -99.524 (0.8172) | -99.808 (0.3493) |
| Day 15 / 3 hrs post-dose | | | |
| n | 9 | 8 | 9 |
| Mean (SD) | -6.693 (65.1702) | -99.562 (0.7422) | -98.306 (4.9029) |
| Day 15 / 6 hrs post-dose | | | |
| n | 9 | 8 | 9 |
| Mean (SD) | 40.340 (104.7373) | -97.808 (5.9400) | -99.884 (0.1039) |
| **Tumor Necrosis Factor-α (ng/L)** | | | |
| Day 1 / pre-dose<br>(Baseline) | | | |
| n | 9 | 8 | 9 |
| Mean (SD) | 0 | 0 | 0 |
| Day 1 / 1 hr post-dose | | | |
| n | 9 | 8 | 9 |
| Mean (SD) | 7.035 (34.0620) | -99.650 (0.8967) | -99.708 (0.3022) |
| Day 1 / 2 hrs post-dose | | | |
| n | 9 | 8 | 9 |
| Mean (SD) | 6.927 (64.5909) | -99.670 (0.5194) | -99.749 (0.1755) |
| Day 1 / 3 hrs post-dose | | | |
| n | 9 | 8 | 9 |
| Mean (SD) | 17.264 (77.0445) | -99.856 (0.1141) | -99.613 (0.6064) |
| Day 1 / 6 hrs post-dose | | | |
| n | 9 | 8 | 9 |
| Mean (SD) | 73.479 (114.4272) | -99.564 (0.7004) | -99.391 (1.2838) |
| Day 15 / pre-dose | | | |

| Visit / Timepoint<br>Statistics | Placebo<br>(N=9) | E6742 100 mg BID<br>(N=8) | E6742 200 mg BID<br>(N=9) |
|---|---|---|---|
| n | 9 | 8 | 9 |
| Mean (SD) | -13.886 (70.0234) | -94.934 (9.0841) | -81.991 (51.9123) |
| Day 15 / 1 hr post-dose | | | |
| N | 9 | 8 | 9 |
| Mean (SD) | 16.311 (86.2430) | -99.513 (0.7714) | -99.752 (0.3110) |
| Day 15 / 2 hrs post-dose | | | |
| n | 9 | 8 | 9 |
| Mean (SD) | 24.837 (99.9210) | -99.882 (0.1546) | -99.843 (0.1576) |
| Day 15 / 3 hrs post-dose | | | |
| n | 9 | 8 | 9 |
| Mean (SD) | 44.159 (124.3807) | -99.937 (0.0966) | -99.500 (1.3060) |
| Day 15 / 6 hrs post-dose | | | |
| n | 9 | 8 | 9 |
| Mean (SD) | 105.575 (189.5408) | -99.775 (0.3418) | -99.812 (0.2987) |

Only subjects with non-missing data at both baseline and the relevant post-baseline visit are included in the change/percent change from baseline summary statistics.

For the purpose of all descriptive statistics the median measured concentration in the analytical sample (n=3) is used. If 1/3 measurements were below the limit of quantification (BLQ), the BLQ was assigned as zero and the median value was used; if 2/3 were BLQ, the lower limit of quantification was used; if all measurements were BLQ, zero was used.

**Supplemental Figure S1 Box plots of changes in QTcF values among systemic lupus erythematosus patients under treatment with E6742.**

The upper plot shows aggregated changes in QTcF values and the lower plot shows changes from baseline after multiple doses of placebo or E6742 (100 or 200 mg) administered twice daily (BID). Whiskers extend to the minimum and maximum values, or 1.5 × the interquartile range. QTcF, corrected QT interval using Fridericia’s formula.

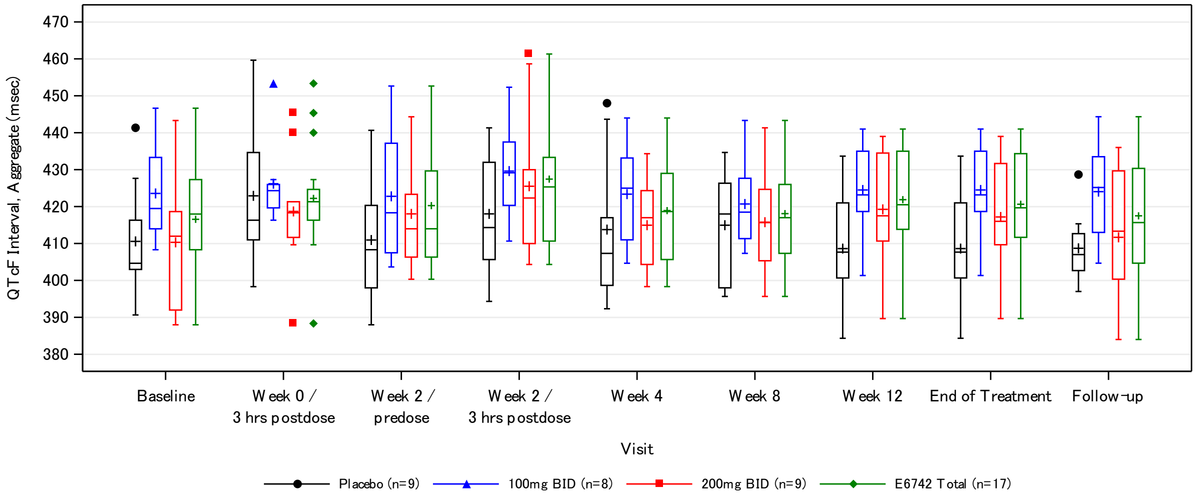


Number of Subjects at Visit

| | Baseline | Week 0 / 3 hrs postdose | Week 2 / predose | Week 2 / 3 hrs postdose | Week 4 | Week 8 | Week 12 | End of Treatment | Follow-up |
|---|---|---|---|---|---|---|---|---|---|
| Placebo | 9 | 9 | 9 | 9 | 9 | 9 | 9 | 9 | 9 |
| 100mg BID | 8 | 8 | 8 | 8 | 8 | 8 | 8 | 8 | 8 |
| 200mg BID | 9 | 9 | 9 | 9 | 9 | 9 | 8 | 9 | 9 |
| E6742 Total | 17 | 17 | 17 | 17 | 17 | 17 | 16 | 17 | 17 |

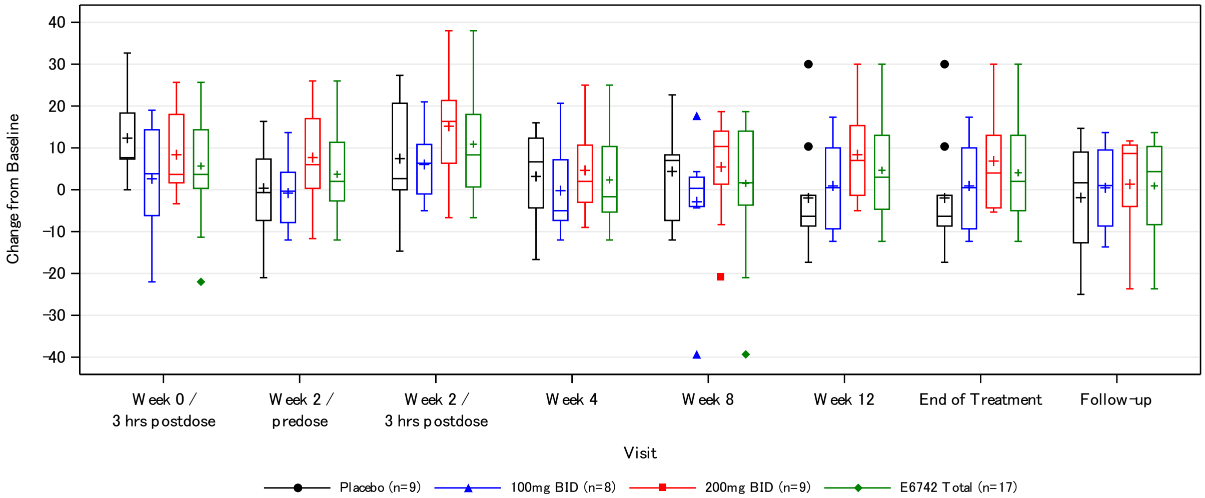


Number of Subjects at Visit

| | Week 0 / 3 hrs postdose | Week 2 / predose | Week 2 / 3 hrs postdose | Week 4 | Week 8 | Week 12 | End of Treatment | Follow-up |
|---|---|---|---|---|---|---|---|---|
| Placebo | 9 | 9 | 9 | 9 | 9 | 9 | 9 | 9 |
| 100mg BID | 8 | 8 | 8 | 8 | 8 | 8 | 8 | 8 |
| 200mg BID | 9 | 9 | 9 | 9 | 9 | 8 | 9 | 9 |
| E6742 Total | 17 | 17 | 17 | 17 | 17 | 16 | 17 | 17 |

**Supplemental Figure S2 Linear-scale plasma concentration-time curve of E6742 in trial patients with SLE.**

Mean (+ standard deviation [SD]) plasma concentrations of E6742 following a single dose (Day 1; upper graph) or multiple doses (Day 15; lower graph) at 100 and 200 mg administered twice daily (BID) in the pharmacokinetic analysis set of patients (N = 17). One patient in the 200 mg BID group did not take the study drug from Days 8–14, and was thus excluded from the data shown on Day 15.

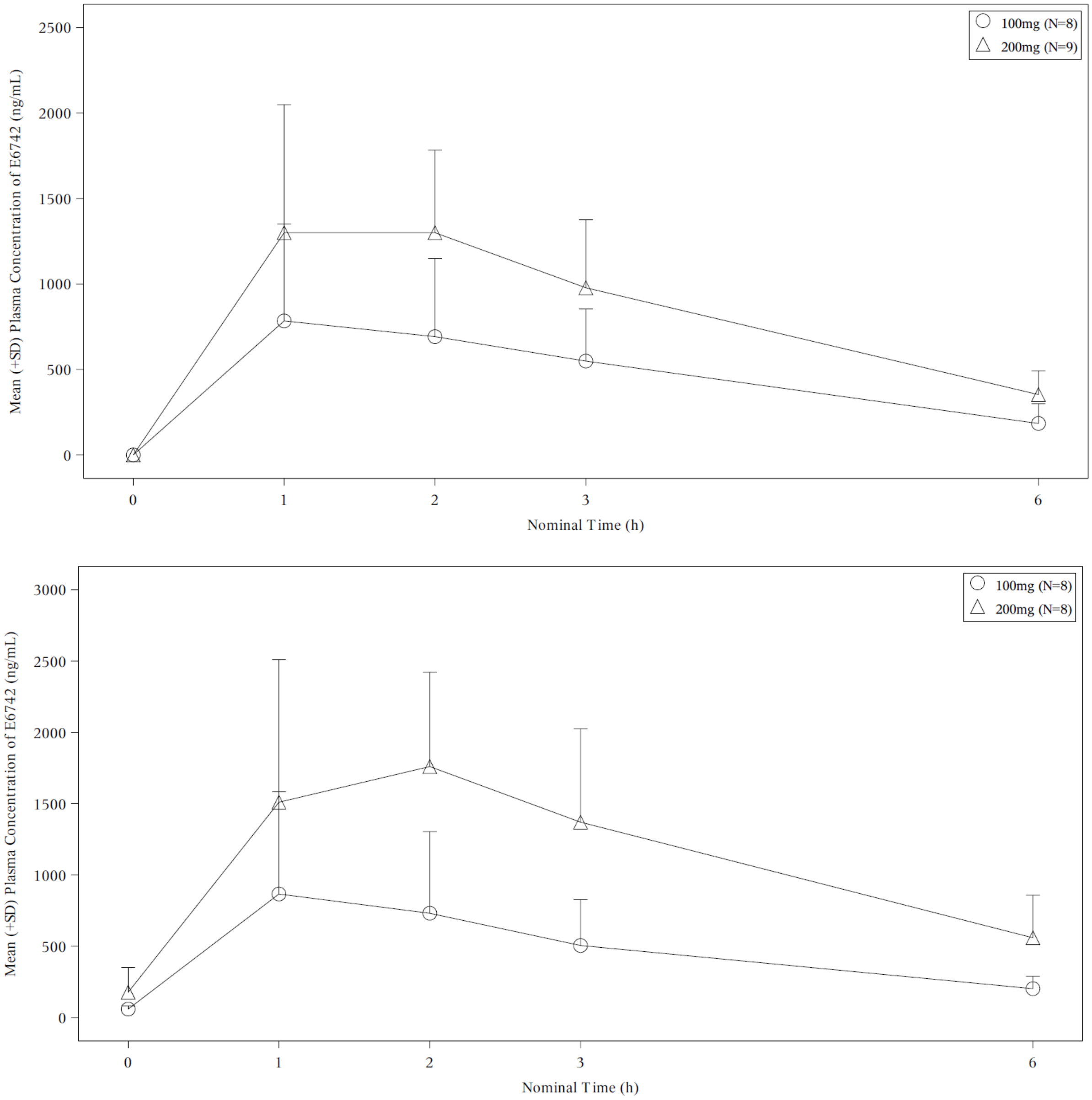
